# A Bayesian Model for Prediction of Rheumatoid Arthritis from Risk Factors

**DOI:** 10.1101/2020.07.09.20150326

**Authors:** Leon Lufkin, Marko Budišić, Sumona Mondal, Shantanu Sur

## Abstract

Rheumatoid arthritis (RA) is a chronic autoimmune disorder that typically manifests as destructive joint inflammation but also affects multiple other organ systems. The pathogenesis of RA is complex where a variety of factors including comorbidities, demographic, and socioeconomic variables are known to influence the incidence and progress of the disease. In this work, we aimed to predict RA from a set of 11 well-known risk factors and their interactions using Bayesian logistic regression. We considered up to third-order interactions between the risk factors and implemented factor analysis of mixed data (FAMD) to account for both the continuous and categorical natures of these variables. The predictive model was further optimized over the area under the receiver operating characteristic curve (AUC) using a genetic algorithm (GA). We use data from the National Health and Nutrition Examination Survey (NHANES). Our optimal predictive model has a smoothed AUC of 0.826 (95% CI: 0.801–0.850) on a validation dataset and 0.805 (95% CI: 0.781–0.829) on a holdout test dataset. Our model identified multiple second- and third-order interactions that demonstrate a strong association with RA, implying the potential role of risk factor interactions in the disease mechanism. Interestingly, we find that the inclusion of higher-order interactions in the model only marginally improves overall predictive ability. Our findings on the contribution of RA risk factors and their interaction on disease prediction could be useful in developing strategies for early diagnosis of RA, thus opening potential avenues for improved patient outcomes and reduced healthcare burden to society.

## 1 Introduction

Rheumatoid arthritis (RA) is a systemic autoimmune disorder of the joints and internal organs that affects 0.5–1.0% of the adult population worldwide^1,2^. It is a major cause of disability and is associated with an increased risk of premature death^3^. The chronic and progressive nature of RA poses a significant financial burden, with the annual societal cost of RA estimated to be $19.3 billion in the United States alone^4^. Despite its profound impact on society and the healthcare system, many aspects of this complex, multifactorial disease remain unknown. A variety of genetic, environmental, and behavioral risk factors have been identified for RA and its association with a number of comorbidities has been reported^2^. Since current medicine does not offer a cure for RA, the major therapeutic goal is preventing the flare-ups, inducing fast remission, and slowing down progressive changes such as irreversible joint deformity^5^. Despite RA’s demand for close and specialized medical supervision, the number of rheumatologists across the United States has been steadily decreasing. There were roughly 5,000 practicing rheumatologists in 2015, but this number is projected to decrease to 3,500 by the year 2025^6^. One promising approach to address this increasing disparity in the patient-to-rheumatologist ratio is the development of analytic tools to facilitate early diagnosis and predict disease progression, thus enabling better access to care and improving the plan for managing the disease.

RA has a strong connection to age and sex. Disease onset is most likely between 50 to 75 years of age^2,7^ and females are affected 2–3 times more than males^2^. Race and ethnicity are also known to influence RA; for example, a lower rate of remission and increased disease activity are reported in African-Americans relative to whites^8^. While the reason for such differences is not completely understood, presence of a “shared epitope” that is highly correlated with RA severity and outcome is suggested to underlie the higher incidence of the disease in certain sub-populations^9,10^. Apart from demographic factors, several socioeconomic and behavioral risk factors are identified for RA. Lower socioeconomic status and less education pose a higher risk of developing the disease^11^ as well as experiencing a poorer prognosis^12^. Increased prevalence of RA is observed among individuals with a history of smoking^13,14^; the risk of RA increases with the intensity of smoking^14–16^.

Presence of comorbidities is extremely common with RA, and they often contribute to worse health outcomes^17,18^. Consistent with the complex, systemic nature of RA, these comorbidities often also affect many systems in the body. Among them are widely prevalent chronic conditions such as cardiovascular disease (CVD) and diabetes, which increase the risk of mortality in RA patients^15,19^. Likewise, hypertension and depression increase the risk of disability^15^. Gout, another disease of joints, has been found to have a higher association with RA^20^. Additionally, RA interferes with the antinociceptive pathway, resulting in enhanced pain perception and leading to a greater risk of sleep problems^21,22^. Several of RA’s comorbidities, such as obesity and depression, demonstrate a bidirectional association with RA, implying their presence elevates the risk of developing RA^13,23^. It is of great clinical interest for physicians and researchers to study the concurrent presence of high Body Mass Index (BMI), depression, and CVD in RA patients as it poses a unique clinical repertoire and has significant consequences on affected individuals. Therefore, careful consideration of comorbidities is important for clinicians working in rheumatology care.

Studies have aimed to predict the occurrence of common diseases like CVD to provide early diagnosis or risk assessment using data mining, machine learning algorithms, and mathematical modeling^24^. While some studies have attempted to predict RA using a similar approach^25,26^, these studies were neither very selective in defining relevant factors for disease prediction nor did consider their interactions. Karlson, et. al.^27^ developed prediction models for RA from a combination of clinical and genetic predictors. The models considered age, sex, and smoking as clinical risk factors and studied eight human leukocyte antigen (HLA) and 14 single nucleotide polymorphism (SNP) alleles associated with seropositive RA as genetic risk factors. Models considering either clinical risk factors alone or both clinical and genetic risk factors were compared for discrimination ability using the receiver operating characteristic (ROC) curve. The models with clinical risk factors alone had areas under the ROC curve (AUC) of 0.57-0.63, while models considering both clinical and genetic risk factors had AUC of 0.66-0.75, indicating an improvement of discrimination ability following the inclusion of genetic risk factors. Chibnik, et. al.^28^ developed a weighted Genetic Risk Score (GRS) from 39 alleles associated with an increased risk of RA. After controlling for age and smoking, the authors used GRS in a logistic regression to discriminate between non-RA and four phenotypes of RA in the NHS dataset. Their model predicted seronegative, seropositive, erosive and seropositive, and erosive RA with AUCs of 0.563, 0.654, 0.644, and 0.712, respectively. Several other studies^29–32^ have performed similar predictive analyses using a combination of environmental and genetic risk factors to create models with good discrimination abilities. The best predictive model we are aware of (as measured by AUC) was developed by Scott, et. al.^33^. In this study, the authors considered age, sex, and 25 HLA and 31 SNP alleles to develop a model with an AUC of 0.857 (95% CI: 0.804–0.910), indicating high discrimination ability.

While previous studies have demonstrated the feasibility of predicting RA from environmental and genetic information, patient genetic data are not readily available in a regular healthcare set-up, thus limiting their practical applicability. In this work, we aimed to develop a predictive model of RA using information commonly available in peripheral health centers or rural infrastructures, such as comorbidities, demographic, socioeconomic, and behavioral factors that are known to associate with RA. We used Bayesian logistic regression to build our model, and considered up to third-order interaction between the variables. Furthermore, to reduce the computational need without compromising predictive accuracy, we implemented FAMD and wrapper methods, which allowed the selection of most important variables for the model.

## 2 Methods

### 2.1 Description of data and preprocessing

Subjects in this study were participants in the National Health and Nutrition Examination Survey (NHANES)^34^, a biannual survey designed to assess the health of the US population administered by the Centers for Disease Control and Prevention. NHANES offers freely accessible detailed health datasets on a sample drawn from the US that is representative of the national population. These datasets provide information on demographic variables, socioeconomic condition (SEC), survey questionnaires, and bio-specimen examinations. Participants are deidentified and represented by a unique sequence number in each dataset.

NHANES data cohorts from 2007 to 2016 were used in this study, providing an initial dataset with 48,484 participants. Information on demographics, medical conditions, depression, body measures, blood pressure, diabetes, smoking habits, and sleep were obtained from each release cycle, giving a total of 11 variables. Data for gender, age, ethnicity, and SEC were obtained from the demographics datasets. SEC was measured using the ratio of a participant’s family’s income to their poverty threshold (IPR). Participants 17 years old or younger were excluded from the analysis to prevent confounding effects from juvenile RA. Participants were divided into five ethnic categories in the ethnicity variable: Mexican-American (MA), other Hispanic (OH), white, black, and other non-Hispanic (ONH). The ethnicity variable was coded into four new dummy variables using the white ethnicity as the reference category because it contained the largest number of participants. Self-reported diagnoses of RA and gout were obtained from the medical questionnaire dataset. Depression was measured using the nine-question Patient Health Questionnaire (PHQ)^35^. Scores on each of the nine questions were manually summed to create a quasi-continuous variable for measuring depression. BMI for each participant was obtained from the body measures dataset as a continuous measurement of obesity. Systolic blood pressure (BP) was calculated from the average of four readings in the blood pressure dataset. Self-reported diagnosis of diabetes were used in this analysis. Borderline diabetes was not considered in the diabetic category. Participants were included in the smoking category if they indicated smoking of at least 100 cigarettes in their life on the smoking questionnaire. Nightly hours of sleep were recorded in one-hour increments with a maximum of 12 to accommodate for variations in NHANES data collection between 2007 to 2014 and 2015 to 2016.

Participants who responded “don’t know,” refused to respond, or had missing data for any variable were excluded from this study. We created second- and third-order interactions between the independent variables by multiplying the initial variables together (with the exception of sequence number and RA). Interactions created by squaring binary variables and multiplying mutually exclusive binary variables were removed from the dataset. New variables that represent an interaction between two or three initial variables are termed “interacted” variables. Quantitative variables were centered and scaled to have means of zero and standard deviations of one. This dataset was further divided into training, validation, and test datasets according to a 50–25–25% split for use in model building and validation.

### 2.2 Factor Analysis of Mixed Data

The added interacted variables are highly-correlated, posing a problem for regression analysis. Using factor analysis of mixed data (FAMD)^36^ new uncorrelated synthetic variables were created, and data projected onto them. FAMD effectively performs principal component analysis (PCA) on quantitative variables and multiple correspondence analysis on qualitative variables. PCA takes in observations of correlated variables and constructs a change of coordinates such that the synthetic output variables are decorrelated. Similarly, multiple correspondence analysis takes in observations of nominal categorical variables and returns a set of decorrelated synthetic variables that represent underlying structures in the original data. In both cases the physical interpretability of the created variables is sacrificed in order to obtain favorable statistical properties, allowing efficient representation of data by a small set of uncorrelated variables.

In FAMD, a new synthetic variable *v* is created by maximizing the criterion

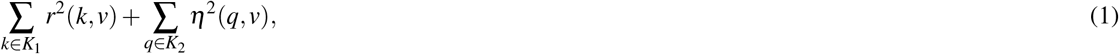

where *K*_1_ are qualitative variables, *K*_2_ are continuous variables, *r*^2^ is Pearson’s correlation statistic, and *η*^2^ is the effect size measure from analysis of variance models^37^. A complete disjunctive coding was performed on all qualitative variables. This created a pair of indicator variables corresponding to each state of every categorical variable in the dataset, all of which were already boolean variables. This process creates *K*_2_ indicator variables that are only used in FAMD. The original categorical variables were kept as supplementary variables in the dataset (variables that are not used for calculating the synthetic variables but are projected onto them for interpretation), while all remaining quantitative and indicator variables are active variables (used for calculating the synthetic variables).

A decorrelated set of synthetic variables maximizing equation (1) can be computed using the singular value decomposition (SVD) of the data matrix ***M***, whose columns correspond variables and rows to observations, that is values of those variables for participants. SVD was performed on all active variables, amounting to calculating matrices ***M*** = ***U* Σ*V***^**T**^, used to project the data onto orthogonal axes (synthetic variables). ***U*** is an orthogonal matrix used to calculate projections of the participants onto the synthetic variables. ***V*** was used to find the projections of the active variables on the new synthetic variables. The projections of the categorical variables onto the synthetic variables are determined from their indicator variables. **Σ** is a diagonal matrix containing the singular values, which are in turn square-roots of variance they explain in the dataset so that **Σ**^2^ is the (diagonal) covariance matrix of the synthetic (decorrelated) variables. Synthetic variables corresponding with variances less than one were omitted to maintain low intercorrelation after the validation and test datasets were projected onto them. Due to properties of SVD, discarding low-variance synthetic variables is known to be optimal approach, in the sense of (1), to construction of reduced-order representation of the data. FAMD was performed using the package `FactoMineR`^38^ in R 3.6.0.

### 2.3 Statistical Analysis

Bayesian logistic regression was used to predict RA in this study. This approach was preferred because of the common-sense statistical interpretations it provides^39^. Bayesian regression summarizes model coefficients and predictions with probability distributions. The results are frequently reported by the highest density interval (HDI) capturing 99% of the probability in the posterior distribution for coefficients. Here 50% and 99% HDIs were included in the interval plots of the posterior distributions. Variables are ranked in the interval plots based on the posterior probabilities that their coefficients are greater or less than one (when transformed from log-odds to odds scale). If a coefficient’s median is greater than one, the probability that it is greater than one is calculated, Pr(*β* > 1 |*y*). However, if a coefficient’s median is less than one, the probability that it is less than one is used, Pr(*β* < 1 |*y*). Ties between coefficients with equal probabilities of being greater or less than one are broken using the absolute values of the medians of their posterior distributions. A Bayesian approach also allows prior information about model coefficients to be specified with a probability distribution, known as the prior distribution. In this study, we specify uniform prior distributions for all model coefficients because the large amount of information in the data causes the likelihood term to dominate when estimating coefficients, effectively making any prior information irrelevant^39^. Markov chains used to sample the posterior distribution were required to have potential scale reduction factors below 1.1 to indicate approximate convergence, imposing a more stringent requirement than the one recommended by Brooks and Gelman^40^. Bayesian logistic regression was performed using `Stan in R 3.6.0` through the package `RStan`^41^.

### 2.4 Predictive Performance and Feature Selection

A wrapper approach to feature selection was implemented in this study to identify the optimal subset of synthetic variables to predict RA. A wrapper approach (as opposed to a filter or embedded approach) uses the predictive performance of subsets of synthetic variables to identify the optimal subset. The predictive performance of the regression models in the genetic algorithm (GA, described below) was determined using the area under the receiver operating characteristic curve. Binormal smoothing of the ROC curve is implemented for its robustness in obtaining an unbiased estimate of the model’s true discrimination ability^42^. This assumes that the distributions of the predicted probabilities of response for the positive and negative cases can be described by a pair of normal distributions, *y*_1_ and *y*_0_, respectively:

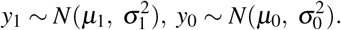

In this study, the binormally smoothed AUC is calculated using two parameters:

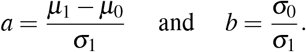

The AUC is calculated as

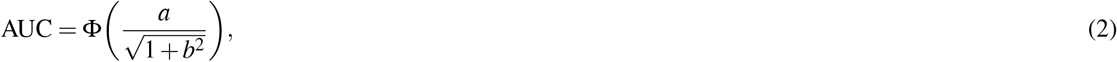

where Φ is the standard normal cumulative distribution function. Estimates for *a* and *b* are obtained by linear regression to the equation

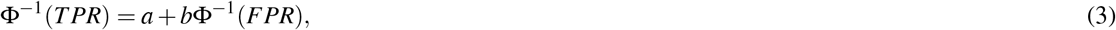

where *TPR* and *FPR* represent the true positive and false positive rates across all thresholds of classification.

A radial sweep is used to generate confidence bands for the ROC curve to provide optimal coverage^43^. Equation (3) is transformed to polar coordinates with center (FPR = 1, TPR = 0) in ROC space. *r* is calculated for values of *θ* in increments of 0.01 from zero to *π*/2. Confidence intervals (CIs) for the AUC and for values of *r* are found from 10,000 bootstrapped samples of the predicted probabilities used to generate the ROC curve.

We used a GA in this study to implement a wrapper approach to feature selection. The GA performs an optimization to find the best subset of synthetic variables for predictive performance according to the AUC (2). The GA was parameterized to have a population size of 500 and run for 150 generations. The GA was seeded with variable subsets always containing the first seven synthetic variables and randomly containing the remaining 45 synthetic variables. All computation for the GA was performed using a server from the Clarkson Open Source Institute at Clarkson University with two Intel Xeon E5-2650 processors with 192 gigabytes of usable physical memory. Running the GA on this server took approximately two weeks.

Rank selection was used to determine which variable subsets would be selected for genetic transformation to create the next population. The probability that a subset *x* would be selected is given by

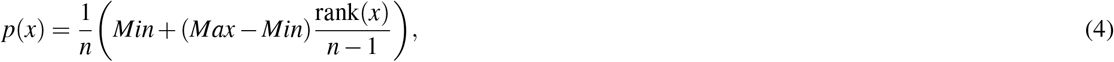

where *n* is the size of the population. *Min* represents the expected number of times the subset with the poorest predictive ability is selected, while *Max* represents the same for the subset with the best predictive ability, with the constraint that *Min* + *Max* = 2 is imposed^44^. rank(*x*) gives the rank of the variable subset within the population such that the best subset has rank *n*. In this study, *Min* was set to 0.7 and *Max* was set to 1.3 to allow for substantial generational improvement while maintaining sufficient exploration of the search space.

Each variable subset had a probability of 0.8 for being selected for single-point crossover, which was used for its simplicity and performance^45^ in GAs. Each subset was also subject to a 0.1 probability of being randomly mutated. Elitism was implemented using 5% of the population to maintain high-quality solutions throughout the GA’s search. The optimal subset was tested on a holdout set of data to assess for overfitting.

### 2.5 Coefficient Reconstruction

HDIs for the coefficients of the original variables are obtained from the posterior distributions of the optimal subset of synthetic variables. The optimal feature subset of size *n* was fit to the data using eight Markov chains each with 400 samples of the posterior distribution, creating a 3200-row by *n*-column matrix ***A*** of the probability distributions of the coefficients for synthetic variables in the logistic regression model. Columns corresponding to synthetic variables that were omitted were set to zero in ***A***. Probability distributions for the coefficients of the interacted variables ***B*** are calculated from ***V*** according to the equation below.

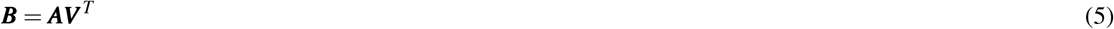

Estimates for the binary variables in the interacted dataset were obtained from the difference between the estimates for their indicator variables.

## 3 Results

### 3.1 Variable Selection

Selection of risk factor variables to incorporate in our model for RA prediction was guided by their reported association with RA and data availability in the NHANES database. These variables include disease comorbidities (diabetes, depression, high BMI, hypertension, and gout), demographic factors (gender and ethnicity), socioeconomic factors (IPR), and behavioral factors (smoking and sleep hours) (Figure 3). Consistent with the literature, the NHANES dataset demonstrated an association of these risk factors with RA, although the extent of the difference varied. For example, RA was less common among males (41.5% of RA subjects. However, the gender disparity was substantially smaller than reported by previous studies (Figure 3**a**)^2^. This difference could be attributed to the survey-based diagnosis of RA, the data preprocessing procedure, and the inherent design of NHANES (Table S1). Subjects with RA were also more likely to suffer from diabetes, gout, high BMI, depression (measured by PHQ score), and high BP (Figure 3**a**,**c**). Risk of RA was found to increase with age, and it was more common among black ethnicity^46^ but substantially less prevalent among the ONH population (Figure 3**b**,**c**). Behavioral factors such as smoking was observed more among RA subjects, while sleep has a less conspicuous impact even though it was reported previously^22^. Interestingly, subjects with RA were found have a lower IPR, suggesting an association of RA with lower economic status.

**Figure 1.**
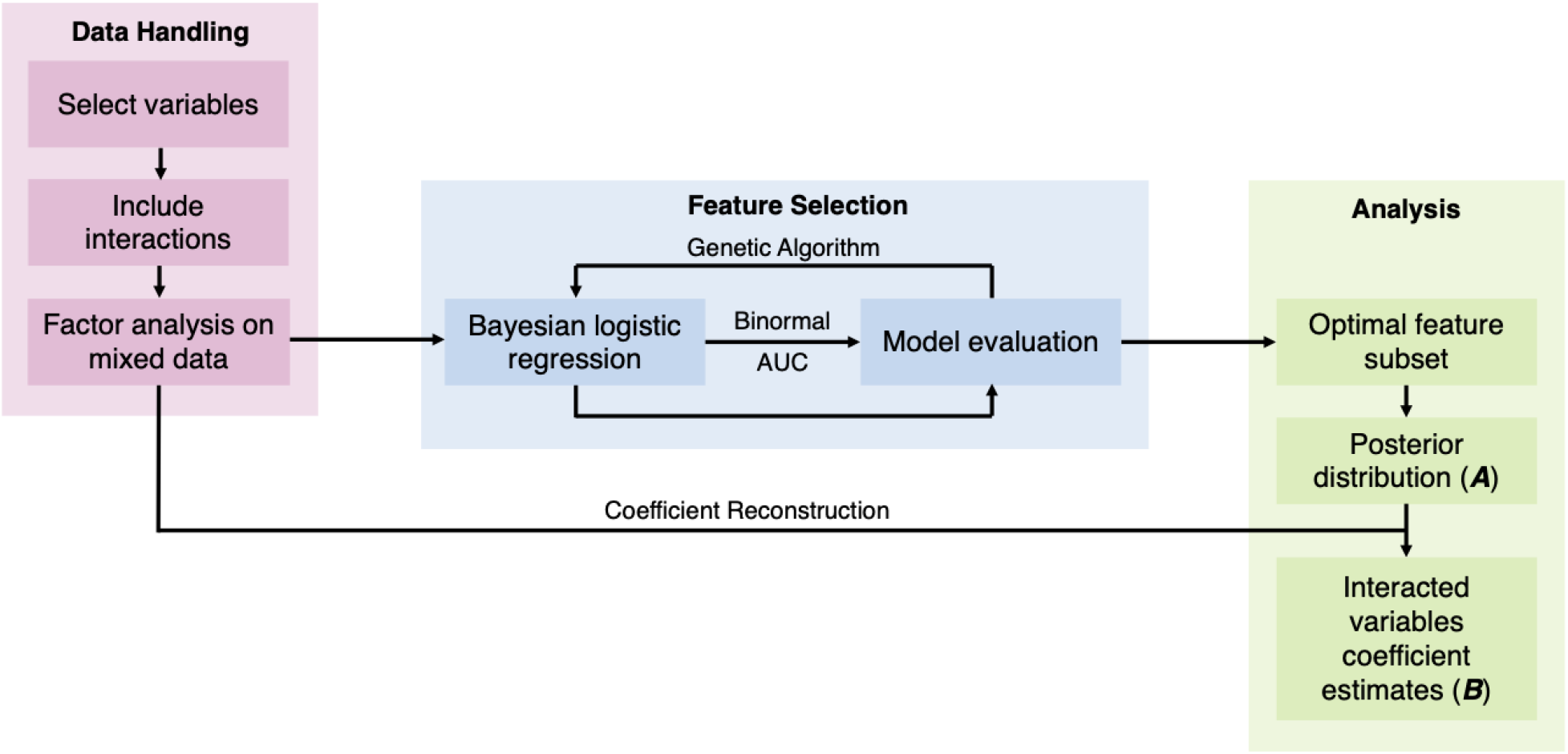
Study methods diagram.

**Figure 2.**
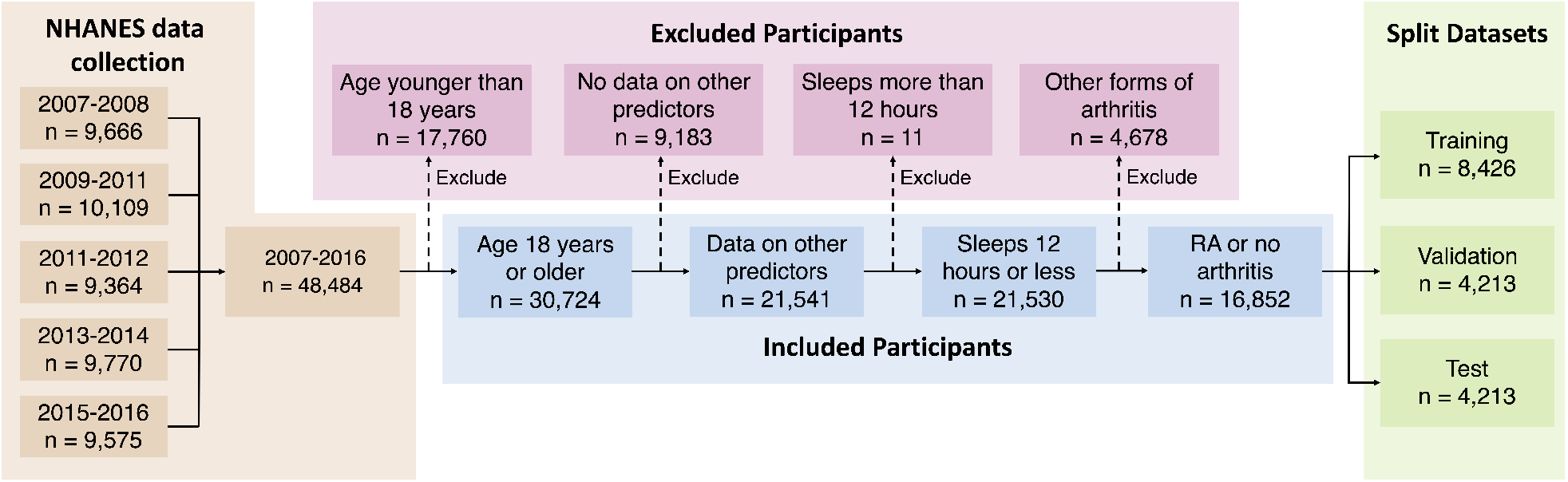
Selection of study population.

**Figure 3.**
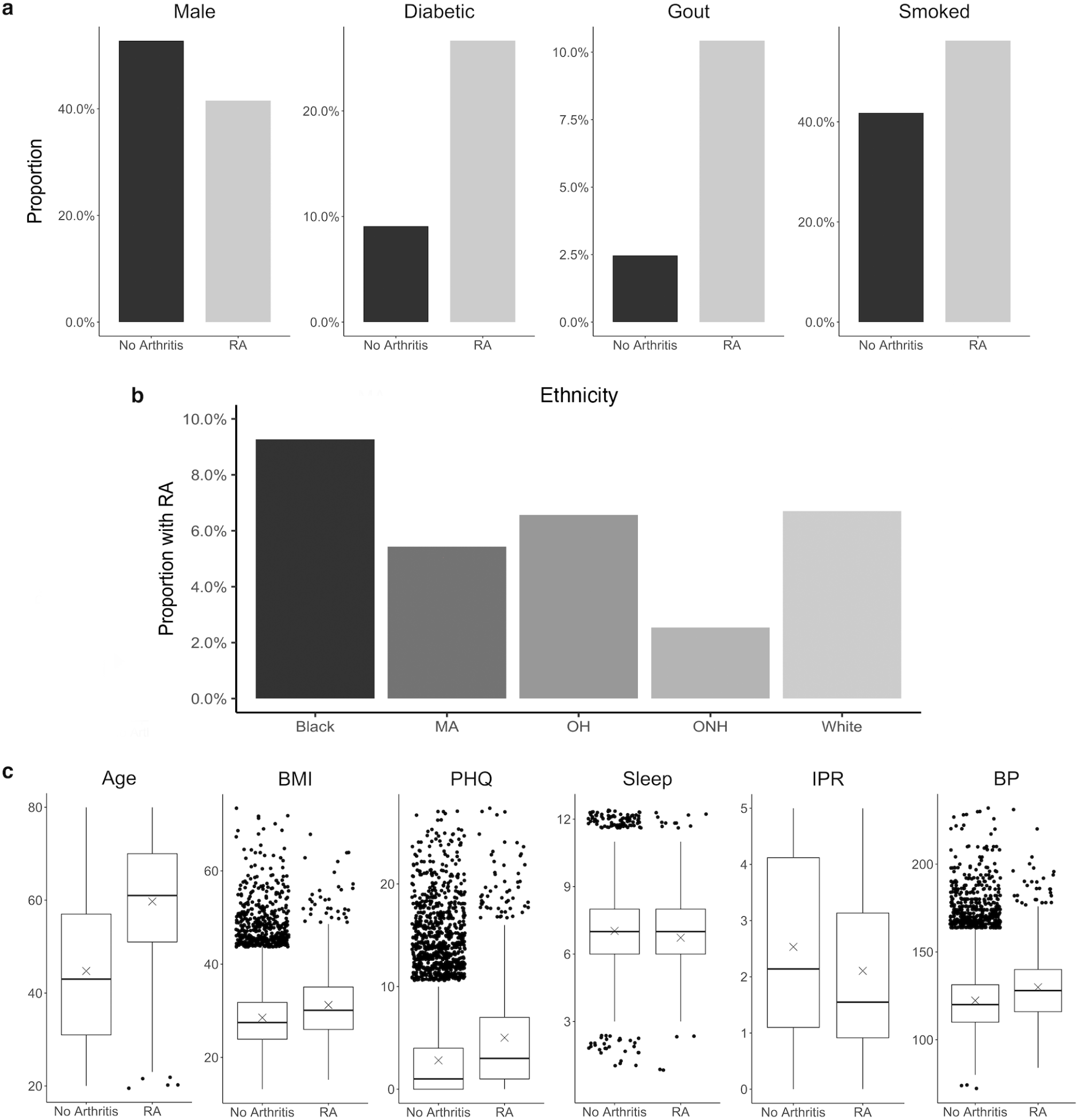
Distribution of various RA risk factors in the study population. (**a**) Comparison between RA and no arthritis population for risk factors coded as binary variables. (**b**) Prevalence of RA among various ethnicity. (**c**) Comparison between RA and no arthritis population for risk factors coded as continuous variables. Units: Age, years; BMI, kg/m^2^; PHQ, PHQ score; Sleep, hours; IPR, nondimensional; Systolic, mmHg. Abbreviations: Mexican-American, MA; other Hispanic, OH

A total of 11 risk factors were considered in our study, which generated 14 first-order variables after including 4 binary variables obtained from dummy-coding ethnicity (using the white population as the reference category). For model building and validation, the dataset was further divided into training, validation, and test categories (Table 1). The distribution of the variables were found to be nearly equivalent across each category, indicating an even split after data preprocessing. A slightly greater variation among the three datasets was observed for the RA group, which could be attributed to a substantially smaller number of individuals in this group than the control no arthritis group. In order to analyze second- and third-order interactions, we created 475 interaction variables from the 14 first-order variables, leading to a total of 489 variables.

**Table 1.** Summary characteristics of all participants, RA, and no arthritis (None) groups in training, validation, and test datasets. Proportions satisfying conditions are reported binary variables. Sample means and standard deviations are reported for continuous variables. Units: Age, years; BMI, kg/m^2^; PHQ, PHQ score; Sleep, hours; IPR, unitless; Systolic, mmHg. Abbreviations: Mexican-American, MA; other Hispanic, OH; other non-Hispanic, ONH.

| Prop. | Training (n = 8,426) |  |  | Validation (n = 4,213) |  |  | Test (n = 4,213) |  |  |
| --- | --- | --- | --- | --- | --- | --- | --- | --- | --- |
|  | All | RA | None | All | RA | None | All | RA | None |
| Male | 0.523 | 0.436 | 0.529 | 0.513 | 0.353 | 0.524 | 0.523 | 0.434 | 0.529 |
| Gout | 0.030 | 0.103 | 0.024 | 0.033 | 0.112 | 0.028 | 0.027 | 0.100 | 0.022 |
| Diabetic | 0.106 | 0.270 | 0.095 | 0.099 | 0.283 | 0.087 | 0.098 | 0.244 | 0.088 |
| Smoked | 0.428 | 0.555 | 0.419 | 0.419 | 0.543 | 0.410 | 0.435 | 0.595 | 0.424 |
| MA | 0.163 | 0.128 | 0.166 | 0.157 | 0.152 | 0.157 | 0.163 | 0.129 | 0.166 |
| OH | 0.105 | 0.097 | 0.106 | 0.114 | 0.123 | 0.114 | 0.102 | 0.111 | 0.101 |
| Black | 0.208 | 0.305 | 0.201 | 0.216 | 0.309 | 0.210 | 0.205 | 0.269 | 0.200 |
| White | 0.410 | 0.427 | 0.408 | 0.402 | 0.368 | 0.404 | 0.412 | 0.448 | 0.409 |
| ONH | 0.114 | 0.043 | 0.119 | 0.111 | 0.048 | 0.115 | 0.118 | 0.043 | 0.124 |
| $\bar{x}$ (s) | | | | | | | | | |
| Age | 45.6 (16.9) | 59.9 (13.3) | 44.6 (16.6) | 45.9 (17.1) | 59.5 (13.4) | 45.0 (16.9) | 45.8 (17.1) | 59.6 (13.2) | 44.8 (16.9) |
| BMI | 28.7 (6.65) | 31.4 (7.49) | 28.5 (6.54) | 28.9 (6.81) | 31.6 (7.75) | 28.7 (6.70) | 28.6 (6.59) | 30.6 (7.91) | 28.4 (6.47) |
| PHQ | 2.89 (4.03) | 4.80 (5.45) | 2.76 (3.88) | 3.01 (4.16) | 5.37 (5.69) | 2.84 (3.98) | 3.03 (4.02) | 5.11 (5.18) | 2.88 (3.88) |
| Sleep | 7.01 (1.44) | 6.83 (1.68) | 7.02 (1.42) | 7.02 (1.47) | 6.79 (1.76) | 7.03 (1.45) | 7.01 (1.44) | 6.46 (1.81) | 7.05 (1.40) |
| IPR | 2.50 (1.64) | 2.13 (1.50) | 2.53 (1.64) | 2.50 (1.63) | 2.05 (1.50) | 2.54 (1.63) | 2.51 (1.65) | 2.11 (1.48) | 2.54 (1.65) |
| BP | 123 (17.9) | 130 (19.9) | 122 (17.6) | 123 (18.0) | 130 (20.6) | 122 (17.7) | 123 (18.3) | 129 (19.7) | 122 (18.1) |

### 3.2 Predictive Performance

To build our model, we first excluded the highly correlated variables from the total set of variables containing higher-order interactions. Since our data contained both categorical and continuous variables, we implemented FAMD to identify the correlated variables. A total of 52 synthetic variables with variances greater than one were obtained by FAMD that represented 92.3% of the variation in the training data. Table 2 summarizes these synthetic variables according to the percentage of variance explained by each of them. A feature selection from these synthetic variables was further performed by a wrapper approach using GA. An optimal subset containing 33 of these synthetic variables was identified that provides the greatest discrimination ability. Figure 4**a** shows the progression of the GA’s search to find the subset of synthetic variables that best predicts RA. 33 of the 52 total synthetic variables were selected through this process, which was able to predict RA with a smoothed AUC of 0.826 with 95% CI of 0.801–0.850 (Figure 4**b**). The potential scale reduction factors 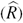 and estimated coefficients from the final regression model for these selected synthetic variables are shown in Table 2. For variables omitted through the feature selection process, the medians for posterior distributions of coefficients (*β*) were set to one and do not have 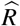 values (Table 2). This subset of variables was also used on the test dataset to obtain a smoothed AUC of 0.805 (95% CI: 0.781–0.829), indicating high accuracy on external data and that the model was not overfitting to the training dataset during regression or the validation dataset during feature selection.

**Table 2.** Percentage of variance explained, 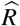, and medians for posterior distributions of coefficients for synthetic variables (*β*) returned by FAMD. Synthetic variables omitted through feature selection have *β* set to one and do not have 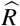 values.

| Var | % Exp | $\hat{R}$ | $\beta$ | Var | % Exp | $\hat{R}$ | $\beta$ | Var | % Exp | $\hat{R}$ | $\beta$ | Var | % Exp | $\hat{R}$ | $\beta$ |
| --- | --- | --- | --- | --- | --- | --- | --- | --- | --- | --- | --- | --- | --- | --- | --- |
| 1 | 10.3 | 1.00 | 1.0963 | 14 | 1.60 | 1.00 | 0.9788 | 27 | 0.849 | — | 1 | 40 | 0.442 | 1.00 | 0.9086 |
| 2 | 7.30 | 1.00 | 1.0545 | 15 | 1.59 | 1.00 | 0.9239 | 28 | 0.821 | — | 1 | 41 | 0.433 | — | 1 |
| 3 | 6.65 | 1.00 | 0.9542 | 16 | 1.49 | 1.00 | 0.8932 | 29 | 0.802 | 1.00 | 1.1096 | 42 | 0.419 | — | 1 |
| 4 | 6.49 | 1.00 | 0.9640 | 17 | 1.24 | — | 1 | 30 | 0.740 | 1.00 | 1.0633 | 43 | 0.387 | — | 1 |
| 5 | 6.17 | 1.00 | 0.9967 | 18 | 1.21 | 1.00 | 0.9688 | 31 | 0.658 | 1.00 | 1.0092 | 44 | 0.367 | — | 1 |
| 6 | 5.96 | 1.00 | 0.9963 | 19 | 1.18 | 1.00 | 0.9598 | 32 | 0.630 | 1.00 | 0.9839 | 45 | 0.343 | — | 1 |
| 7 | 5.19 | 1.00 | 0.9836 | 20 | 1.15 | 1.00 | 1.0240 | 33 | 0.590 | 1.00 | 0.9812 | 46 | 0.336 | — | 1 |
| 8 | 4.26 | 1.00 | 1.0169 | 21 | 1.07 | — | 1 | 34 | 0.552 | — | 1 | 47 | 0.304 | 1.00 | 0.9828 |
| 9 | 3.31 | 1.00 | 0.8923 | 22 | 1.04 | — | 1 | 35 | 0.537 | 1.00 | 0.9777 | 48 | 0.281 | — | 1 |
| 10 | 2.91 | 1.00 | 1.0987 | 23 | 1.02 | — | 1 | 36 | 0.493 | 1.00 | 1.0614 | 49 | 0.277 | 1.00 | 0.9798 |
| 11 | 2.61 | 1.00 | 0.9484 | 24 | 0.931 | — | 1 | 37 | 0.491 | — | 1 | 50 | 0.262 | 1.00 | 1.0314 |
| 12 | 1.83 | 1.00 | 1.0846 | 25 | 0.903 | 1.00 | 1.0062 | 38 | 0.470 | 1.00 | 0.9688 | 51 | 0.248 | 1.00 | 0.8923 |
| 13 | 1.61 | 1.00 | 1.0362 | 26 | 0.872 | — | 1 | 39 | 0.466 | — | 1 | 52 | 0.230 | — | 1 |

**Table 3.** Summary of key findings for (**a**) first-order, (**b**) second-order, and (**c**) third-order variables. 99% HDIs are shown for estimated regression coefficients on the odds scale.

|  |  |  |  |
| --- | --- | --- | --- |
| (a) | **Risk factor** | $\hat{\beta}$ | **Comments** |
|  | Age | 1.0533–1.0765 | • Aging increases risk for RA<br>• Most influential first-order effect |
|  | Sleep | 0.9526–0.9768 | • More sleep decreases risk of RA |
|  | BMI | 1.0088–1.0308 | • Higher BMI increases risk of RA<br>• Weaker first-order effect |
|  | BP | 0.9856–1.0153 | • No direct effect on risk of RA |
|  | ONH | 0.9785–1.0032 | • No direct effect on risk of RA |
| (b) | **Risk factor** | $\hat{\beta}$ | **Comments** |
|  | Age<sup>2</sup> | 1.0527–1.0752 | • Effect of increased age on RA risk is greater at older ages |
|  | Age·BMI | 1.0535–1.0770 | • Age and BMI have the strongest second-order interaction |
|  | Male·ONH | 0.9171–0.9793 | • Being male and ONH ethnicity markedly reduces risk for RA |
|  | Age·BP | 1.0421–1.0592 | • Interaction of age and BP increases risk of RA |
|  | BP·Sleep | 0.9585–0.9824 | • Higher BP further lowers RA risk afforded by sleep |
| (c) | **Risk factor** | $\hat{\beta}$ | **Comments** |
|  | Age<sup>2</sup>·BMI | 1.0562–1.0787 | • Strong third-order interaction between age and BMI |
|  | Age<sup>3</sup> | 1.0511–1.0735 | • Effect of increased age on RA risk is greater at older ages |
|  | Age<sup>2</sup>·BP | 1.0459–1.0645 | • Third-order interaction between age and high BP increases RA risk |
|  | Age·BMI·BP | 1.0454–1.0621 | • High BP further increases RA risk from aging and high BMI |
|  | Male·ONH·Sleep | 0.9319–0.9816 | • Sleep adds to lowering RA risk afforded by being male and ONH |

**Figure 4.**
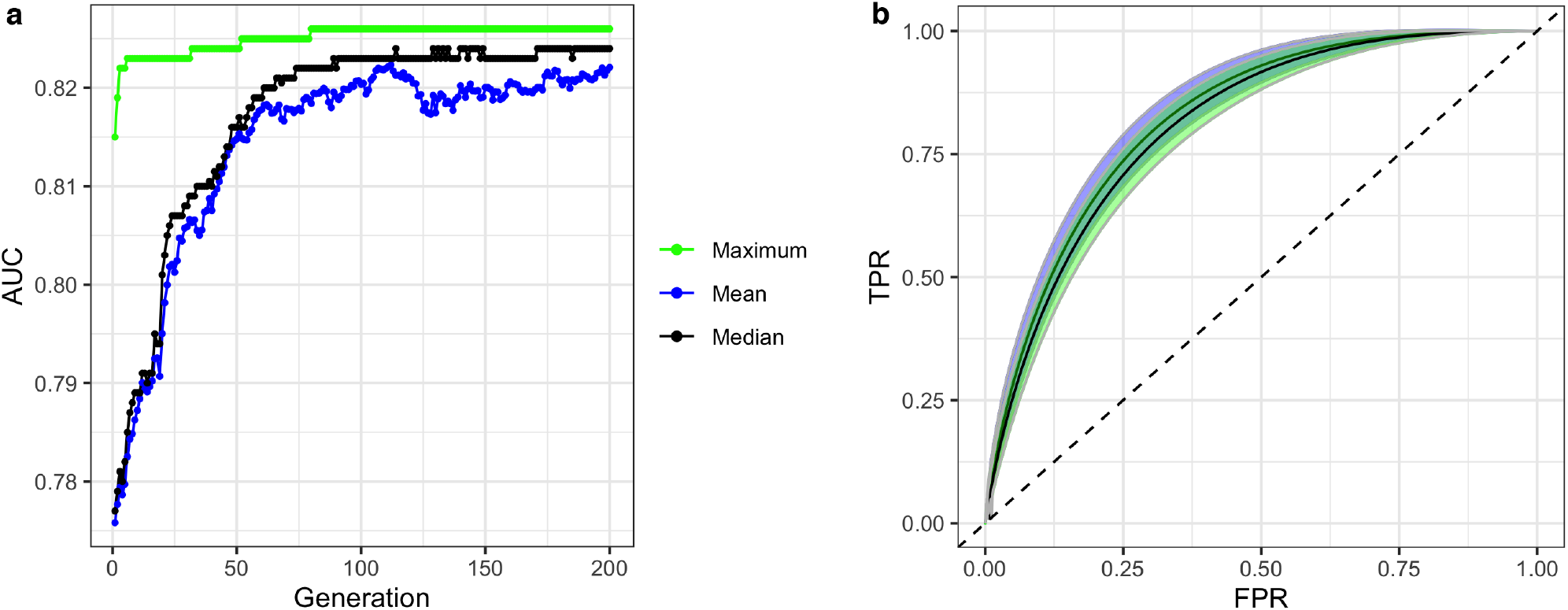
(**a**) convergence of genetic algorithm (GA) on an optimal subset of synthetic variables with maximum, mean, and median fitness values in each generation of the search. (**b**) ROC curves and confidence bands of optimal model predicting on validation datasets (confidence region shaded blue) and test datasets (confidence region shaded green). Dashed line represents the ROC curve of a model with no predictive ability, corresponding to an AUC of 0.5.

Interestingly, we find that even the first-order variables alone are highly predictive, with an AUC of 0.823, and that higher-order interactions yield only a small improvement of AUC to 0.826. Furthermore, our approach can generate a predictive accuracy higher than most previous works reported even when using a small set of first-order variables (Table S2)^27–29^. For example, considering age and smoking alone can generate a model with an AUC of 0.748, and including sex further increased the AUC to 0.772. While these findings suggest the potential of model building from first-order variables alone, future studies are required to identify optimum set of variables that would contribute maximum to the predictive accuracy.

### 3.3 Risk Factor Interactions

The subset of synthetic variables returned by the GA is not easily interpretable on its own. Each synthetic variable represents a latent variable that is a linear combination of the total pool of 489 variables. The posterior distribution of the synthetic variables obtained through this process was used to construct HDIs for each of the 489 variables using equation (5). Furthermore, to allow intuitive comparison across variable types and effect orders, the coefficient estimates were computed for the standardized versions of the variables (Figure 5). Thus, the variables with HDIs further away from 1.0 are more significant predictors of RA, while a narrower interval indicates a greater certainty about how a specific variable affects RA. The analysis aims to identify the effects of first-order variables, and the influence of any second- and third-order interactions as illustrated in Figure 5**a**.

**Figure 5.**
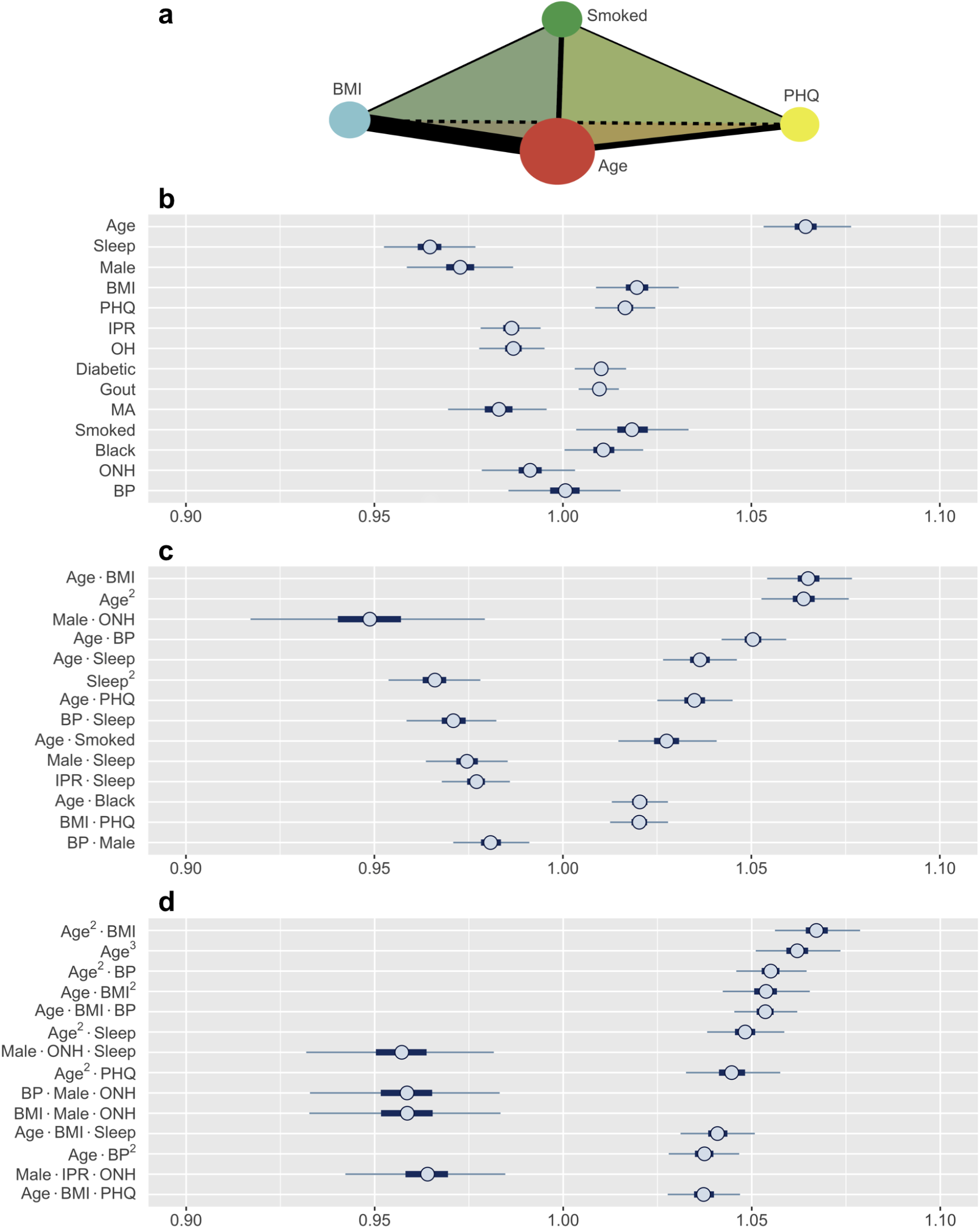
(**a**) A schematic illustrating the interaction analysis between four risk factors. Size of vertices, thickness of edges, and color of faces denote the size of first-, second-, and third-order effects, respectively. (**b-d**) Posterior distributions of standardized coefficients for selected variables. HDIs for (**b**) all first-order variables, (**c**) most influential second-order, and (**d**) third-order variables (inner bounds, 50% HDIs; outer bounds, 99% HDIs). Horizontal scale represents the odds multipliers for risk of RA for one standard deviation increase in value of the variables. Abbreviations: Mexican-American, MA; other Hispanic; OH; other non-Hispanic, ONH.

The prediction of RA in the test dataset by the first-order variables overall aligns well with the association of these variables to RA observed in Figure 3. Age, BMI, depression (PHQ score), diabetes, gout, and smoking are found to be positive predictors, while male gender and financial wellness (IPR) reduce the risk of having RA (Figure 5**b**). A clear influence of ethnicity is also observed: Risk of RA is higher among black population and lower among Mexican-American population when compared against white. Interestingly, sleep emerged as a strong negative predictor (most influential first-order variable after age), even though an association of RA and sleep was not clearly observed in the data. In contrast, systolic BP played no effect on RA prediction as a first-order variable (HDI roughly symmetric about one), although RA subjects had a higher mean systolic BP than the control population.

Apart from the effects of individual first-order variables, we were interested to identify any influence of higher-order interactions in RA prediction. Figure 5**c** enumerates the 14 most influential second-order variables observed in our study. Age turns out to not only be the strongest first-order predictor variable but also to have prominent second-order interactions with several other variables, including BMI, BP, depression, sleep, and smoking. The strongest second-order interaction effect was found between age and BMI (median: 1.0648, 99% HDI: 1.0535–1.0770), which is comparable to the influence of age (1.0642, 1.0533–1.0757) or three times the influence of BMI (1.0196, 1.0088–1.0306), considered individually. Interestingly, the second-order effect of age (1.0636, 1.0527–1.0752) is similar in magnitude to its first-order effect, suggesting that the effect of age on RA risk increases with age. We also observed several second-order interactions to reduce the risk of RA. For example the combination of ONH ethnicity with male gender strongly reduces the risk of having RA (0.9485, 0.9156–0.9797), even though ONH does not have a significant influence in lowering RA risk and male gender has a less prominent effect. This finding suggest the second-order interaction with male gender could underlie low RA prevalence observed among ONH ethnicity (Figure 3**b**). Sleep demonstrates an interesting interaction effect on RA. While increased sleep hours was found to lower the risk of RA, its second-order effect with age increased the risk significantly, suggesting an altered role of sleep on the body’s immune system with aging.

Our model was also able to reveal the existence of strong third-order interactions. Figure 5**d** lists 14 most prominent third-order interactions where we find the frequent appearance of a few variables, with age and BMI being most common. Other factors involved in strong third-order interactions are gender, ONH ethnicity, sleep, depression, and BP. Similar to the second-order interactions, these third-order interactions are seen to either increase or decrease the risk of RA. In particular, for interactions posing high risk, we often observe age and BMI, either as a third-order variant of their interaction between, or in combination with a third variable such as BP, sleep, or depression. By contrast, the coexistence of ONH ethnicity with male gender in a third-order interaction prominently reduces the risk of RA when associated with sleep, BMI, BP, or IPR as the third variable. Thus, variables such as sleep or BP, when involved in third-order interactions, can both increase or decrease the risk of RA, suggesting a complex interplay of underlying physiological mechanisms.

#### Range of Interactions: Age vs. BMI

The finding of several prominent second- and third-order interactions in our model further motivated us to investigate the range of interactions for an individual risk factor. To this direction, we focused on comparing age and BMI, two variables that demonstrated the strongest higher-order interaction (Figure 6). Our analysis shows that these two variables have very different interaction profiles. Age demonstrates strong second-order interactions with multiple comorbidities (BMI, BP, and depression), sleep, and smoking, all of which increase the risk of RA (Figure 6**a**). In contrast, second-order interaction effects to BMI are moderate to weak (except with age) and, depending on the interacting variable, increases or decreases the RA risk (Figure 6**b**). The third-order interactions for age and BMI follow a similar pattern as observed in the second-order interactions, except the combination of male and ONH ethnicity reduces RA risk (Figure 6**c**,**d**). We hypothesize that general changes in body physiology accompanied with aging cause other risk factors to have a greater impact on RA, resulting in these interaction effects. In contrast, high BMI potentially elicits specific influence in the pathophysiology of interacting risk factors, increasing or decreasing the magnitude of the effects. Together, these results confirm that the interactions of a risk factor with other risk factors are highly specific in nature, and are dependent on the variables considered.

**Figure 6.**
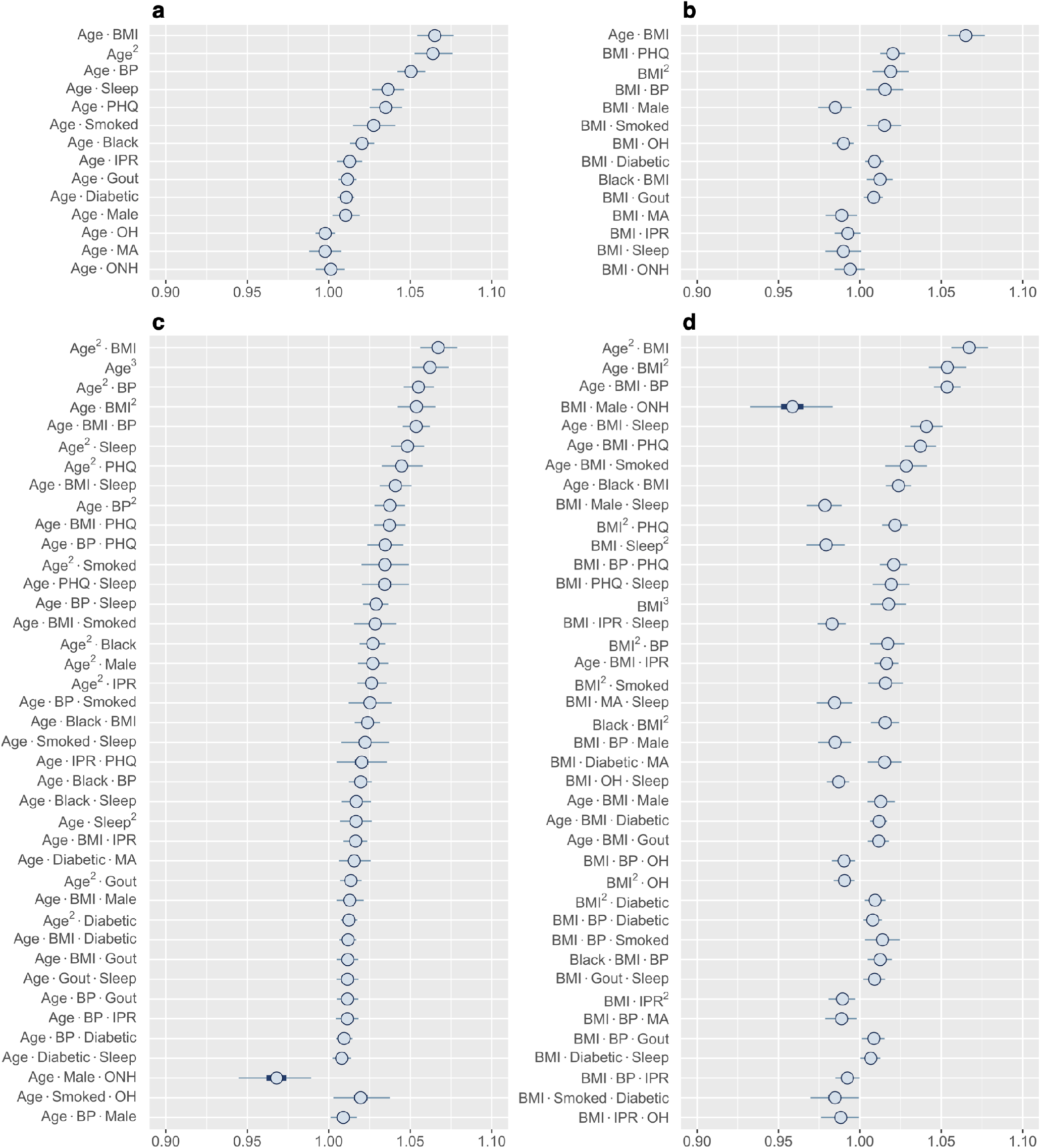
Higher-order interactions of age and BMI. (**a-b**) Posterior distribution of standardized coefficients for all second-order interactions of age (**a**) and BMI (**b**). (**c-d**) Posterior distribution of standardized coefficients for a selection of 40 third-order interactions involving age (**c**) and BMI (**d**). Inner bounds and outer bounds represent 50% HDIs and 99% HDIs, respectively. Abbreviations: Mexican-American, MA; other Hispanic, OH; other non-Hispanic, ONH. Horizontal scale shows odds multipliers for risk of RA for a one standard deviation increase in value of variable.

#### Influence Through Interactions: BP and ONH Ethnicity

Finally, we wanted to explore the higher-order interactions for risk factors that did not show a significant first-order effect. Among all first-order effects, only BP and ONH category had 99% HDIs that contained one (Figure 5**b**). Identifying the most influential second-order interactions for BP or ONH category reveals that 12 out of the top 13 involve BP (Figure 7**a**). The only interaction involving ONH category included in this list (it was also the strongest interaction) is with male gender, strongly lowering the risk for RA. In contrast, the posterior distribution of the interactions of BP indicate that the risk could both increase or decrease depending on the specific interaction. For example, the risk can increase from interaction with age, depression, and BMI, while sleep and male gender reduce the risk. Interestingly, we found the interaction effects of BP with individual risk factors to be similar to their first-order effects. Thus, high BP is expected to enhance the effect of an interaction between risk factors on RA risk. The third-order interactions corroborate well to the second-order interactions with BP occupying 29 of the top 30 interactions (Figure 7**b**). The effect follows the pattern demonstrated by the interaction between the other two factors. While hypertension is generally considered as a comorbidity of RA, there is a lack of consensus on the true association between RA and hypertension^47^. Our finding that BP does not have a significant first-order effect but has prominent interaction effects with coexisting conditions, offers a potential explanation for the varying results reported in the literature.

**Figure 7.**
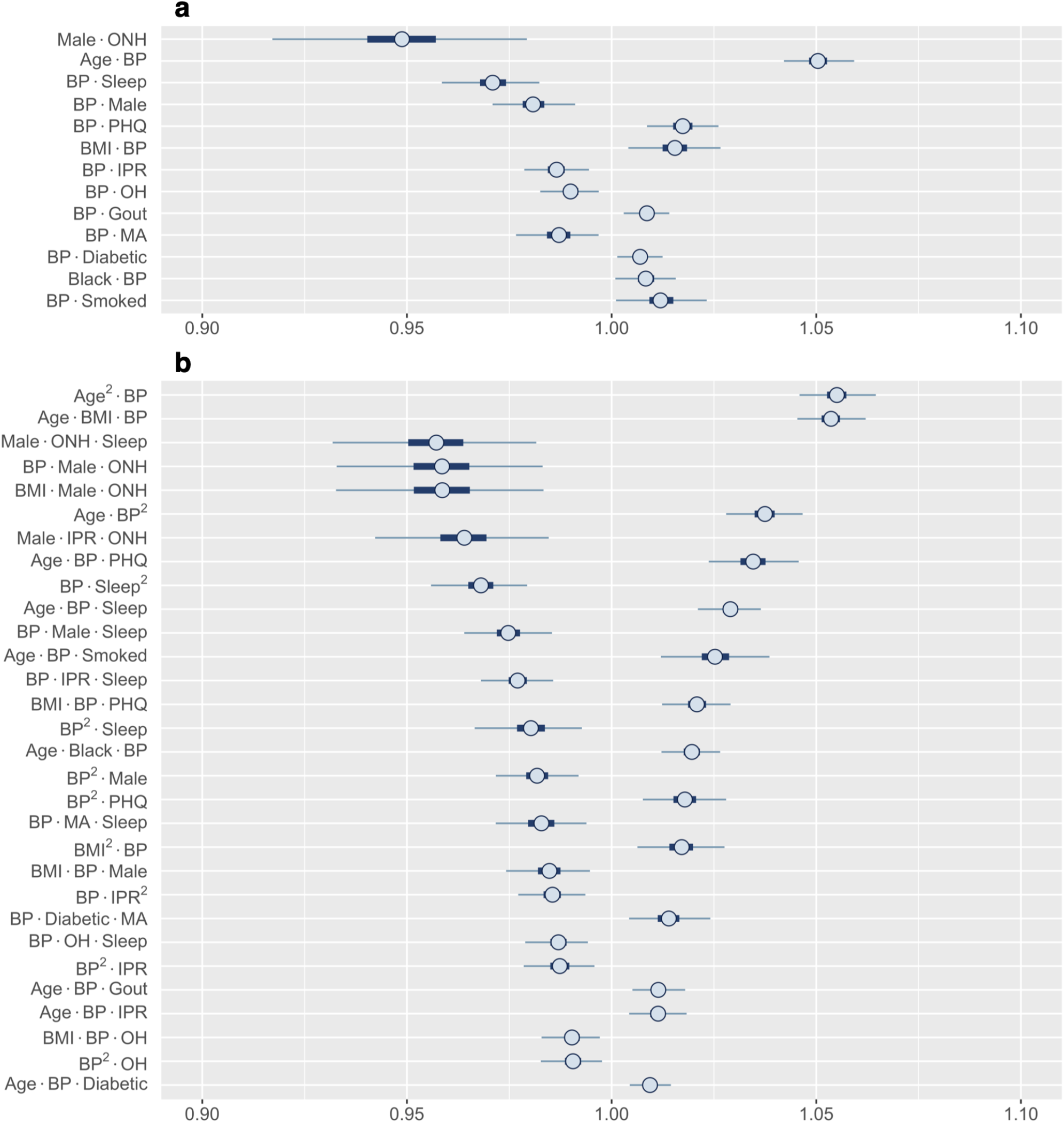
Posterior distributions of most relevant interactions that include either BP or ONH ethnicity as one risk factor. (**a**) 13 most influential second-order interactions and (**b**) 30 most influential third-order interactions are shown. Inner bounds represent 50% HDIs and outer bounds represent 99% HDIs. Standardized coefficient estimates are shown. Abbreviations: Mexican-American, MA; other Hispanic, OH; other non-Hispanic, ONH. Horizontal scale shows odds multipliers for risk of RA for a one standard deviation increase in value of variable.

## 4 Discussion

In this work, we have developed a Bayesian regression model to characterize the risk of RA from common comorbidities, demographic, socioeconomic, and behavioral factors that are known to associate with RA. Apart from providing high predictive accuracy, our model is able to capture the effects of individual variables as well as the important higher-order interactions between them. Consistent with previous literature, known RA risk factors such as depression, high BMI, and smoking are also found to be predictors of RA in our model. Additionally, our model shows that age is not only a key predictor for RA, but also has strong interaction effects with several other variables; prominent among them are BMI, BP, depression, and smoking. Interestingly, some variables such as ONH ethnicity have weak influence as a single-order variable but their combination with certain other variables (male gender in case of ONH ethnicity) could elicit a prominent higher-order interaction. The knowledge of these strong interactions will help to determine if a person is at a higher or lower risk of RA when both conditions coexist.

One of our primary objectives in this study was to identify and elucidate the effects of important higher-order interactions between risk factors in the prediction of RA. The main challenge in performing such a study comes from the exponential increase in the number of synthetic variables as more higher-order interactions are considered, correspondingly increasing the computational cost. This limitation led us to restrict our study to a maximum of third-order interactions. Our implementation of FAMD further reduced the number of predictor variables analyzed during regression, substantially lowering the requirement for computation. FAMD also allowed the consideration of both categorical and continuous risk factor variables in the model.

In our model, we used feature selection to select an optimal subset of synthetic variables. This step was introduced to not only improve the model’s predictive ability but also to obtain a greater precision in determining the effect of risk factors on RA. When studying the manifold interactions between these risk factors, increased precision from feature selection helps to address increases in posterior variances resulting from dramatic increases in the number of variables being analyzed (see Supplementary Fig. S1). We implemented a wrapper method for feature selection. However, there are alternative approaches,the most common being filter methods^48^. Filter methods employ a ranking system to determine the most relevant variables before any prediction is performed^49^, some examples of which include the Pearson correlation coefficient, Fisher score, and mutual information^48^. Filter-based approaches generally perform faster than wrapper methods since they do not require the predictive model to be run simultaneously. However, because of this, they do not necessarily return the optimal subset of features for prediction^49^. Additionally, some filter methods are prone to selecting redundant features^49^, while wrapper methods find the optimal subset based on their performance in the predictive model and do not encounter this issue. Thus, employing a wrapper approach for feature selection allowed us to determine the most important subset of synthetic variables for prediction, and subsequently enabled more precise estimates of the effects of interactions between risk factors on RA. One downside of wrapper methods is that they are generally more computationally expensive than filter methods and implementation of techniques based on exhaustive searches can become computationally infeasible for large datasets^49^. To overcome this limitation, we implement a wrapper approach using a GA, a type of evolutionary algorithm, and is capable of providing high-quality solutions with reasonable computational effort^50^.

The promise of our model in predicting RA is demonstrated by a higher predictive accuracy in comparison to previous studies when only a small number of first-order variables are considered (Table S2). Surprisingly, we find that accounting for up to third-order interactions makes only a small increase in the AUC compared to just first-order effects (0.826 vs. 0.823) even though a strong higher-order interaction between several variables is observed. We speculate that a conflation of RA with other forms of arthritis in NHANES datasets could prevent our model achieving substantially higher predictive abilities after incorporation of higher-order effects. This conflation potentially results from self-reported diagnosis of RA and other arthritis in NHANES, and is reflected by a higher proportion of RA in the population than expected from existing literature^2^ (Table S1).

Our results suggest that high predictive accuracy could be achieved from the first-order model alone when an appropriate set of risk factor variables are selected. While predictive performance might not improve significantly by incorporating higher-order interactions in such a scenario, identifying strong interactions could provide important clinical insight. Furthermore, in situations where health resources are highly constrained with severely limited data availability, higher-order interactions could play a significant role in achieving a sufficient degree of predictive accuracy. Our model could also be applicable to predict other chronic diseases that multiple, potentially interacting, factors are known to be associated with.

Even though NHANES provides a rich dataset of risk factors associated with RA, one limitation of the study comes from the self-reported nature of RA diagnosis. Therefore, the model could be made more robust by future validation and optimization with patient data where more rigorous criteria for RA diagnosis, such as the one provided by the American College of Rheumatology (ACR), is used^51^. The second limitation comes from the cross-sectional nature of the NHANES data, where the old and new RA cases cannot be discriminated. This restricts our model prediction results to be better interpreted as correlation rather than causation of RA. We expect the model accuracy to improve along with the ability to infer a causal relationship on training with longitudinal data where diagnosis of RA can be studied against a population with existing risk factors.

In summary, we have developed a model to predict RA from comorbidities, demographic, socioeconomic, and behavioral risk factors. The model demonstrated a high predictive accuracy in comparison with other models reported in the literature. Moreover, our model was able to identify important second- and third-order interactions between the risk factors, which may have important clinical relevance and stimulate further research to understand the mechanisms underlying such interactions. Since the model prediction utilizes patient information commonly available in a regular healthcare set-up, it has the future potential for translation to the clinical setting.

## Data Availability

Datasets used in this work are publicly available for download from NHANES website

https://wwwn.cdc.gov/nchs/nhanes/

## 5 Acknowledgments

We thank Daniel Fuller for his generous assistance and meaningful suggestions. We also thank the Clarkson Open Source Institute at Clarkson University for access to their servers. We especially thank Graham Northup for his continued help in resolving technical issues.

## 6 Author contributions statement

L.L., S.M. and S.S. conceptualized the study. L.L. developed and executed the procedure. S.M. and M.B. verified the procedure. All authors contributed in writing the manuscript.

## 8 Data availability

Raw datasets are publicly available for download from NHANES website (https://wwwn.cdc.gov/nchs/nhanes/). Codes for analysis will be available upon request.

## 9 Additional Information

### Supplementary Information

**Table S1.**
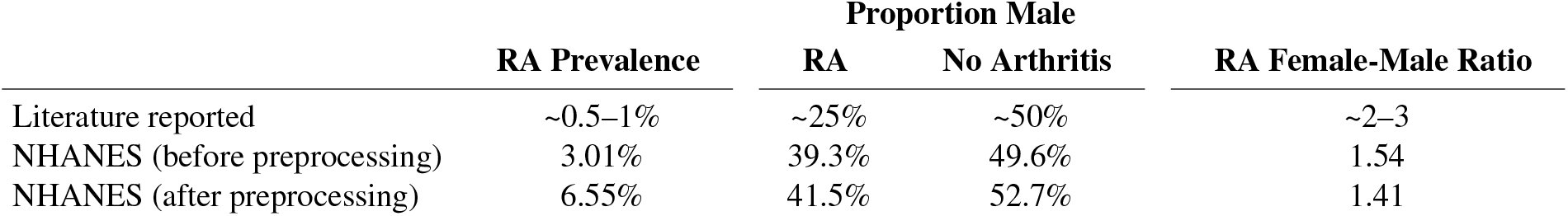
RA prevalence, proportions of males in RA and no arthritis population, and female to male ratio in RA population are compared between literature reported values and NHANES data (before and after preprocessing).

|  | RA Prevalence | Proportion Male |  | RA Female-Male Ratio |
| --- | --- | --- | --- | --- |
|  |  | RA | No Arthritis |  |
| Literature reported | ~0.5–1% | ~25% | ~50% | ~2–3 |
| NHANES (before preprocessing) | 3.01% | 39.3% | 49.6% | 1.54 |
| NHANES (after preprocessing) | 6.55% | 41.5% | 52.7% | 1.41 |

**Table S2.**
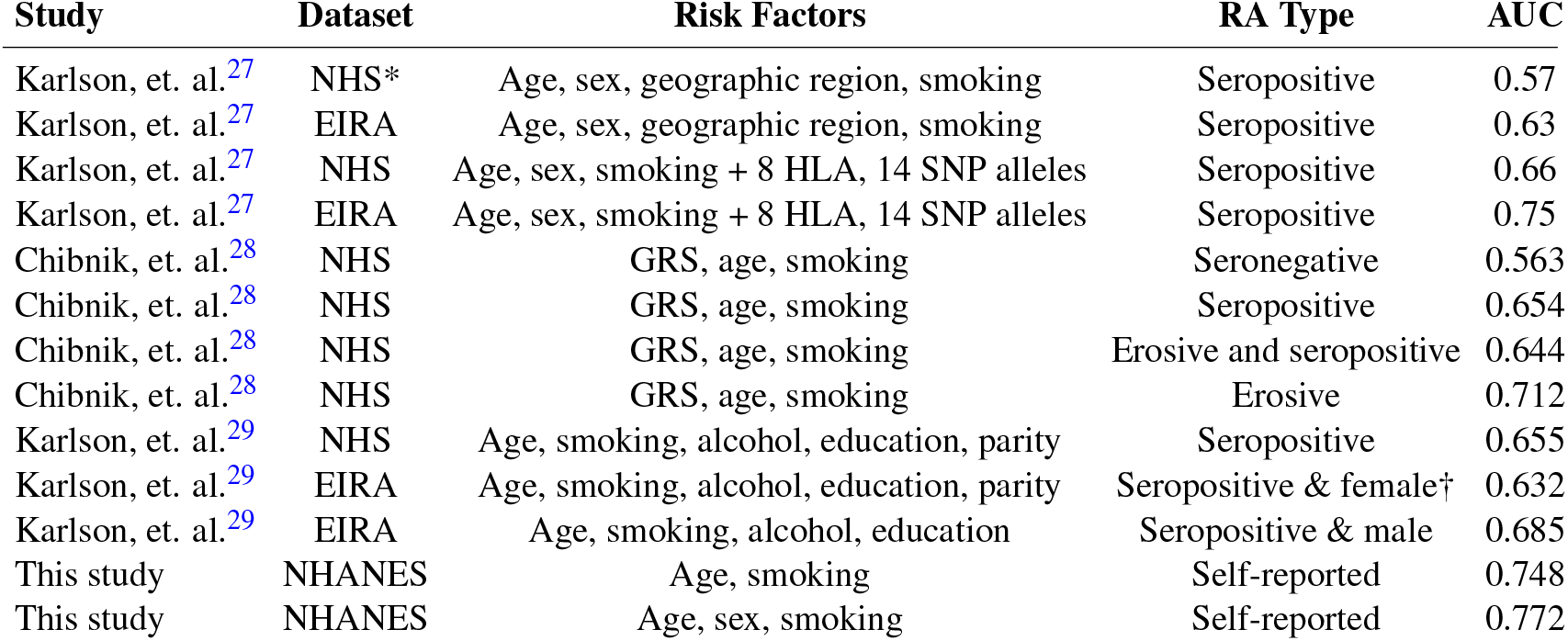
Comparison of predictive ability of current study against from previous works. Abbreviations: NHS, US Nurses’ Health Studies I & II; EIRA, Swedish Epidemiologic Investigation of RA; GRS, Genetic Risk Score. *All prediction using NHS was only performed on females. †”female” indicates prediction was only performed on female subjects; “male” indicates prediction was only performed on male subjects.

**Figure S1.**
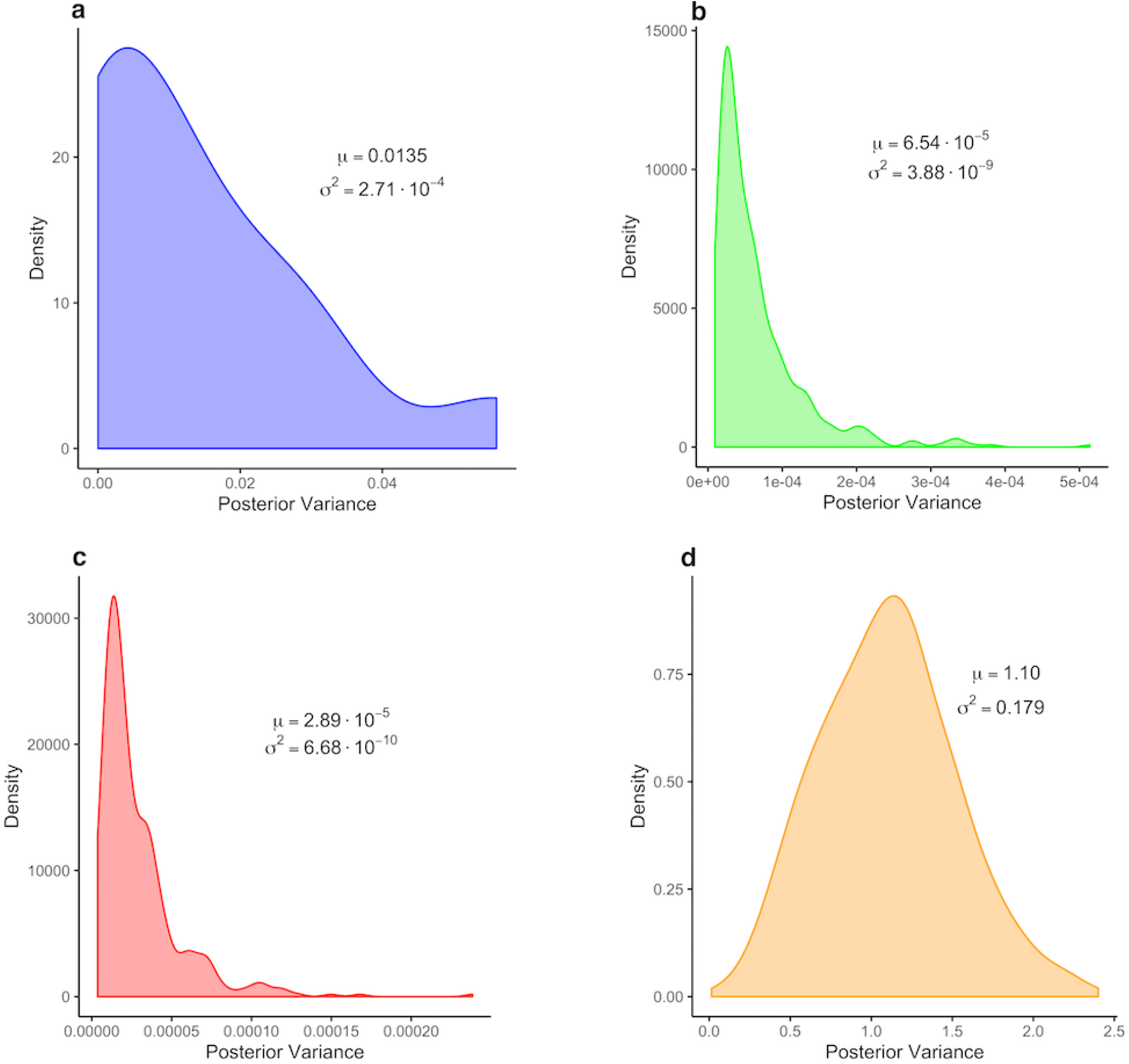
Distributions of posterior variances of estimated regression coefficients from four Bayesian approaches used to predict RA with the training dataset; mean and variance of distributions are shown. (**a**) Regression model using only the 14 first-order variables. (**b**) Estimates for all 489 variables using regression coefficients derived from 52 synthetic variables after FAMD. (**c**) Estimates for all 489 variables using regression coefficients derived from 33 synthetic variables selected by GA. (**d**) regression model using all 489 variables without FAMD.

